# Identifying the Key Predictors of Occupational Fatigue among Long-Distance Truck Drivers in East Africa: A LASSO-Regularized Regression Approach

**DOI:** 10.64898/2026.02.15.26346342

**Authors:** Nelson Kilimo, Kellen Karimi, Olipher Makwaga, Verena Struckmann

## Abstract

**Objectives:** This study aimed to determine the prevalence of occupational fatigue and identify its primary risk factors among long-distance truck drivers operating along the key Kenya-Uganda transport corridor, a vital artery for regional commerce where comprehensive data has been limited.

**Methods:** A cross-sectional analytical study was conducted with 207 exclusively male long-distance truck drivers at the Busia and Malaba border points. Participants completed structured questionnaires capturing demographics, work patterns, sleep habits, and stimulant use. Fatigue was assessed using the Chalder Fatigue Scale. Data were analyzed in RStudio, using LASSO regression with 10-fold cross-validation for predictor selection to address multicollinearity. Selected variables were used in a multivariable logistic model to calculate adjusted odds ratios (aORs). Bootstrap validation assessed the model’s performance.

**Results:** The overall prevalence of occupational fatigue was 51.7%. Key risk factors identified included high pressure to meet deadlines, the use of stimulants (e.g., caffeine, khat) to maintain alertness, and excessively long average shift lengths. The multivariable model demonstrated excellent and stable performance, with a mean Area Under the ROC Curve (AUC) of 0.987 across 1,000 bootstrap samples.

**Conclusions:** Occupational fatigue is highly prevalent among long-distance truck drivers in this region, driven largely by organizational factors. The findings highlight an urgent need for multimodal interventions, including enforceable regulations on driving hours, targeted driver education, and improved scheduling practices by transport companies to safeguard driver well-being and public road safety.

**Key Messages:** *What is already known on this topic:* Truck-driver fatigue is a global problem, but evidence from East African corridors has been minimal.

*What this study adds:* Fatigue prevalence on the Kenya–Uganda corridor is high (51.7%). LASSO analysis highlights key predictors: deadline pressure, stimulant use, long shifts, and poor rest.

*How this study might affect research, practice or policy:* Findings support driving-hour regulations, improved scheduling by transport companies, and targeted public health messaging on stimulant risks.

## Introduction

Driver fatigue is widely recognized as a critical global public health and occupational safety concern, particularly within the transport and logistics sectors(Arnold et al., 1997; Belzer & Sedo, 2018; Casey et al., 2024). Fatigue impairs vigilance, reaction time, and decision-making capacity factors essential to safe driving performance. (Al-Mekhlafi, Isha, Chileshe, Abdulrab, Kineber, et al., 2021; Al-Mekhlafi, Isha, Chileshe, Abdulrab, Saeed, et al., 2021; Dorrian et al., 2006) found that driving fatigue partially or fully mediates the relationship between work factors and driving performance, highlighting its critical role in accident prevention. Notably, such fatigue-related crashes are about 50% more likely to result in death or severe injury because drivers who fall asleep at the wheel typically fail to brake or take evasive action before impact(Ahlström & Anund, 2025; Akyurek et al., 2025; Bener et al., 2017). Similar patterns have been observed internationally, emphasizing that fatigue-induced impairment constitutes a major determinant of crash severity and frequency(Friswell & Williamson, 2013; Philip et al., 2019).

Occupational fatigue, often described as a persistent state of physical or mental exhaustion that reduces a worker’s ability to perform safely and efficiently, arises from prolonged working hours, irregular rest periods, monotonous tasks, and extended wakefulness (Hasan et al., 2022). Within the context of the transport industry, fatigue is both a physiological and psychological hazard. Physiologically, fatigue manifests through sleep deprivation, circadian rhythm disruption, and physical strain caused by factors such as prolonged sitting, vibration, exposure to noise, and temperature extremes. Psychologically, fatigue emerges from job stressors such as long hours, social isolation, high time pressure, and limited autonomy (Jenni et al., 2019). These combined conditions make truck driving one of the most physically and mentally demanding occupations globally (Friswell & Williamson, 2019; Rudin-Brown & Filtness, 2023).

Globally, low- and middle-income countries (LMICs) bear a disproportionate burden of road traffic injuries and fatalities, accounting for nearly 93% of global road deaths despite having only about 60% of the world’s vehicles (World Health Organization, 2023). In these settings, fatigue-related crashes among professional drivers are exacerbated by limited enforcement of occupational health standards, long and irregular driving schedules, and poor road infrastructure (Amoadu et al., 2023). Understanding how fatigue develops and interacts with individual, occupational, and environmental factors is therefore crucial for developing effective interventions to enhance road safety and occupational well-being.

In South Africa, the number of heavy commercial vehicles (HCVs) involved in fatal crashes rose dramatically by over 109% between 2009 and 2011 (Amoadu et al., 2023; Baulk & Fletcher, 2012; Chan, 2011; Maldonado et al., 2002), underscoring the urgent need for fatigue management interventions within the trucking industry. Similarly, in East Africa and particularly in Kenya driver fatigue has been officially recognized as a leading contributor to road crashes. The Kenya Road Safety Policy by the State Department for Public Service, 2023 emphasizes fatigue management as a central strategy for reducing crash risk, recommending structured rest breaks (15-30 minutes every 200 km), maximum daily driving limits (8-10 hours per 24-hour period), and advance scheduling of transport requests to facilitate adequate rest and planning. These policy measures align with international best practices but face implementation challenges due to limited compliance monitoring and inconsistent enforcement mechanisms.

Empirical evidence demonstrates that between 25% and 45% of commercial vehicle drivers globally experience work-related fatigue (Amoadu et al., 2023). In China, for instance, 24.8% of long-distance truck drivers reported fatigue, with higher prevalence observed among less experienced drivers, those operating under tight delivery schedules, and those engaging in unsafe practices such as speeding and overloading (Chen & Zhang, 2016). Similarly, studies across Europe and Latin America indicate that long working hours, night driving, and irregular sleep cycles are strongly associated with fatigue-related crashes(Al-Mekhlafi, Isha, Chileshe, Abdulrab, Saeed, et al., 2021; Barck-Holst et al., 2021; Girotto et al., 2015).

For long-distance drivers, fatigue represents not merely a personal health risk but also a systemic operational hazard that threatens logistics efficiency and public safety. Extended driving hours, limited rest opportunities, and prolonged waiting times for cargo loading and unloading significantly heighten fatigue risk(Friswell & Williamson, 2013). The consequences are twofold: reduced driver alertness increases the likelihood of crashes, and the resulting accidents impose substantial economic costs through loss of cargo, vehicle damage, and human casualties (Montoro et al., 2022; Soliani et al., 2023).

In the East African context, these risks are amplified along major international transport corridors such as the Northern Corridor, a vital artery linking the port of Mombasa with Uganda, Rwanda, and other landlocked countries. The corridor facilitates the majority of regional freight movement, and its continuous operation depends on the physical and psychological well-being of truck drivers who often traverse long distances under intense time pressure and limited rest facilities. Despite the sector’s economic importance, there is limited research examining occupational fatigue and its determinants among drivers in this region.

This study therefore investigated the prevalence and risk factors of occupational fatigue among long-distance truck drivers operating at the Kenya-Uganda border points of Busia and Malaba. By analyzing work patterns, rest practices, and coping mechanisms among this population, the study seeks to elucidate the underlying contributors to fatigue and inform the development of targeted interventions to improve both road safety and occupational health outcomes. Understanding the interplay between individual, occupational, and environmental determinants of fatigue within this high-demand corridor is crucial for reducing fatigue-related crashes and promoting sustainable transport systems in East Africa.

## 2. Methodology

### 2.1 Site selection

This study was conducted among long-distance truck drivers operating along the Kenya-Uganda transport corridor, specifically at the Busia and Malaba border towns, which form part of the Northern Corridor linking the Port of Mombasa to inland East Africa. These sites were strategically selected due to their high volume of freight traffic, representing a critical cross-section of regional logistics operations. The approximately 930-kilometre route from Mombasa to the borders provided an ideal context for assessing fatigue in drivers traveling extended distances under demanding road conditions. The study employed a cross-sectional analytic design with a quantitative approach, using a structured questionnaire to collect data on socio-demographic factors, work patterns, rest duration, fatigue levels, and coping mechanisms. This design was suitable for estimating the prevalence of occupational fatigue and identifying associated predictors within a single observation period.

### 2.2 Study Population, Eligibility, and Sampling Strategy

The study population comprised all long-distance truck drivers departing from Kenya through Busia and Malaba after originating from Mombasa, regardless of nationality.

Eligible participants were those aged 28 years and above, holding valid NTSA class C or E driver’s licenses, and with at least four years of long-distance driving experience. Drivers on medication affecting sleep or alertness were excluded to avoid confounding. The operational definition of occupational fatigue was the presence of at least three symptoms, including excessive daytime sleepiness, difficulty concentrating, physical exhaustion, and reduced alertness. The sample size of 207 drivers was calculated using the (Charan & Biswas, 2013) formula for comparing two proportions, assuming 95% confidence, 80% power, and an odds ratio of 2.0 for the association between inadequate rest and fatigue (Rodríguez Del Águila & González-Ramírez, 2014). A systematic random sampling technique was used: the first truck nearest to customs clearance was enrolled, and thereafter every fourth truck driver was selected.

The waiting period during customs clearance (approximately 25 minutes) provided a practical window for recruitment and survey completion, ensuring representativeness across both border sites.

### 2.3 Variables, Measurement, and Data Collection Procedures

The primary outcome, occupational fatigue, was measured using the validated Chalder Fatigue Questionnaire (CFQ-11), which captures both physical and mental fatigue through 11 Likert-scaled items(Chalder et al., 1993; Jackson, 2015). Scores were dichotomized using the binary method, where a total of four or more indicated fatigue. Other explanatory variables included age, education, marital status, health status, financial strain, and social support.

Data were collected by trained research assistants using structured, interviewer-administered questionnaires translated into Kiswahili for clarity. All assistants underwent standardized training to minimize interviewer bias, ensure consistency, and uphold participant confidentiality.

### 2.4 Data Processing, Statistical Analysis, and Quality Control

Data were collected using Google forms, and then exported as CSV file for analysis in RStudio (version 4.3.2). Descriptive analyses summarized continuous variables using means and standard deviations or medians and interquartile ranges, and categorical variables using frequencies and percentages. The prevalence of fatigue was estimated with 95% confidence intervals. Group comparisons used independent t-tests for continuous variables and chi-square tests for categorical variables.

In recent years, various statistical and machine learning models including logistic regression, gradient boosting machines (GBM), neural networks, XGBoost, Support Vector Machines (SVM), Random Forest (RF), and K-nearest neighbor (KNN) have been widely applied in predictive health research(El_Rahman, 2021; Faul et al., 2009; Li et al., 2022; Omondiagbe et al., 2019). However, these methods can have limitations, such as a propensity for over-fitting, convergence to local minima, and suboptimal performance with imbalanced datasets. The least absolute shrinkage and selection operator (LASSO) is a regression analysis method that addresses these issues by applying a penalty function to select the most relevant predictive features from a larger set of variables. Predictors with nonzero coefficients are retained to construct a more robust and interpretable model(Abd-elnaby et al., 2022; Li et al., 2022).

The aim of this study was to establish and validate a multivariable prediction model to identify drivers at high risk of occupational fatigue. The model was developed using variables selected by LASSO logistic regression. Based on the feature selection performed by LASSO, we established a parsimonious prediction model and developed a nomogram to visualize the results. This tool is designed to help identify high-risk individuals proactively, enabling targeted intervention programs for driver safety and well-being while avoiding unnecessary burden on low-risk populations.

Predictor screening began with univariable logistic regression, retaining all variables with p < 0.2 for further modeling. To address potential multicollinearity and overfitting, a Least Absolute Shrinkage and Selection Operator (LASSO) logistic regression was performed using the *glmnet* package. The optimal penalty parameter (λ) was selected through 10-fold cross-validation, with λ *SE* chosen for parsimony(Abd-elnaby et al., 2022). Variables retained from LASSO were entered into a multivariable logistic regression model to estimate adjusted odds ratios (aORs) and 95% confidence intervals. Model discrimination was assessed via the area under the ROC curve (AUC), and calibration was visually examined. Internal validation was performed using an 80/20 train-test data split with sensitivity, specificity, and confusion matrices computed through the *caret* and *pROC* packages. To ensure data integrity, double data entry verification was conducted, and discrepancies were resolved against original questionnaires(Robin et al., 2011). By employing LASSO regularization, standardized data collection, and validation tests, we minimized analytic and measurement bias and ensured the validity and generalizability of the results.

### 2.5 Ethical Considerations, Limitations, and Mitigation Measures

This study adhered to international ethical standards for research involving human participants. Informed consent was obtained after explaining the study objectives, potential risks, and participants’ right to withdraw without consequence. Ethical approval was obtained from the Kenyatta National Hospital-University of Nairobi Ethics and Research Committee (KNH-UON ERC) (Approval No. P555/07/2024), and research authorization was granted by the National Commission for Science, Technology and Innovation (NACOSTI) (Permit No. NACOSTI/P24/414364).

Administrative notification was made at the Kenya National Highways Authority (KENHA), Customs Offices at Busia and Malaba, and the Busia County Government. The study acknowledged several limitations. The use of self-reported data introduced possible recall and response bias, while the cross-sectional design restricted causal inference. Operational constraints at border posts occasionally limited interview depth.

To mitigate these issues, validated instruments (CFQ-11) enhanced measurement accuracy, and statistical safeguards particularly the LASSO-logistic regression framework minimized model overfitting and confounding. These steps improved both internal validity and interpretability of findings.

## 3. Findings

This chapter highlights the findings of the statistical analyses conducted to identify the key predictors of fatigue among long-distance truck drivers in East Africa. The presentation shows a summary of the predictor selection process using LASSO regression, which identified two significant predictors: Substance Use and High Deadline Pressure. Subsequently, the results of the multivariable logistic regression model are detailed, revealing that substance use and high deadline pressure independently and strongly associated with higher odds of experiencing occupational fatigue. The final model demonstrated excellent discriminatory power, with an Area under the Curve of 0.98, and maintained robust performance upon internal validation using bootstraping.

The study participants comprised 207 exclusively male long distance truck drivers. Demographic analysis revealed a predominantly middle-aged population, with the largest proportion (45.4%) being 40-49 years old, followed by the 30-39 age group (32.4%). The vast majorities of participants were married (91.8%) and had attained a secondary education level (61.8%). Occupational characteristics indicated that most drivers (84.5%) were engaged in full-time employment. Salary distribution was varied, with 36.7% falling into the second quartile (Q2). A significant majority of drivers (91.8%) reported having three or more dependents. Key work practice findings identified concerning patterns.

A high prevalence (84.5%) of drivers reported operating a vehicle for more than four hours without a break. Concurrently, poor record-keeping was common, with 81.2% not maintaining work/rest records. Furthermore, more than half of the surveyed group (53.6%) reported frequently experiencing deadline-related pressure, highlighting a high-stress occupational environment. These findings describe a profile of a predominantly middle-aged, married male workforce operating under significant occupational demands and potential safety risks.

The results of the descriptive and univariable analyses, which provided the foundation for this modeling, are presented in Table 1 and Table 2. To address potential multicollinearity and overfitting, a Least Absolute Shrinkage and Selection Operator (LASSO) logistic regression was performed on a set of nine candidate variables. The optimal penalty parameter was selected via 10-fold cross-validation to yield a parsimonious model.

**Table 1:**
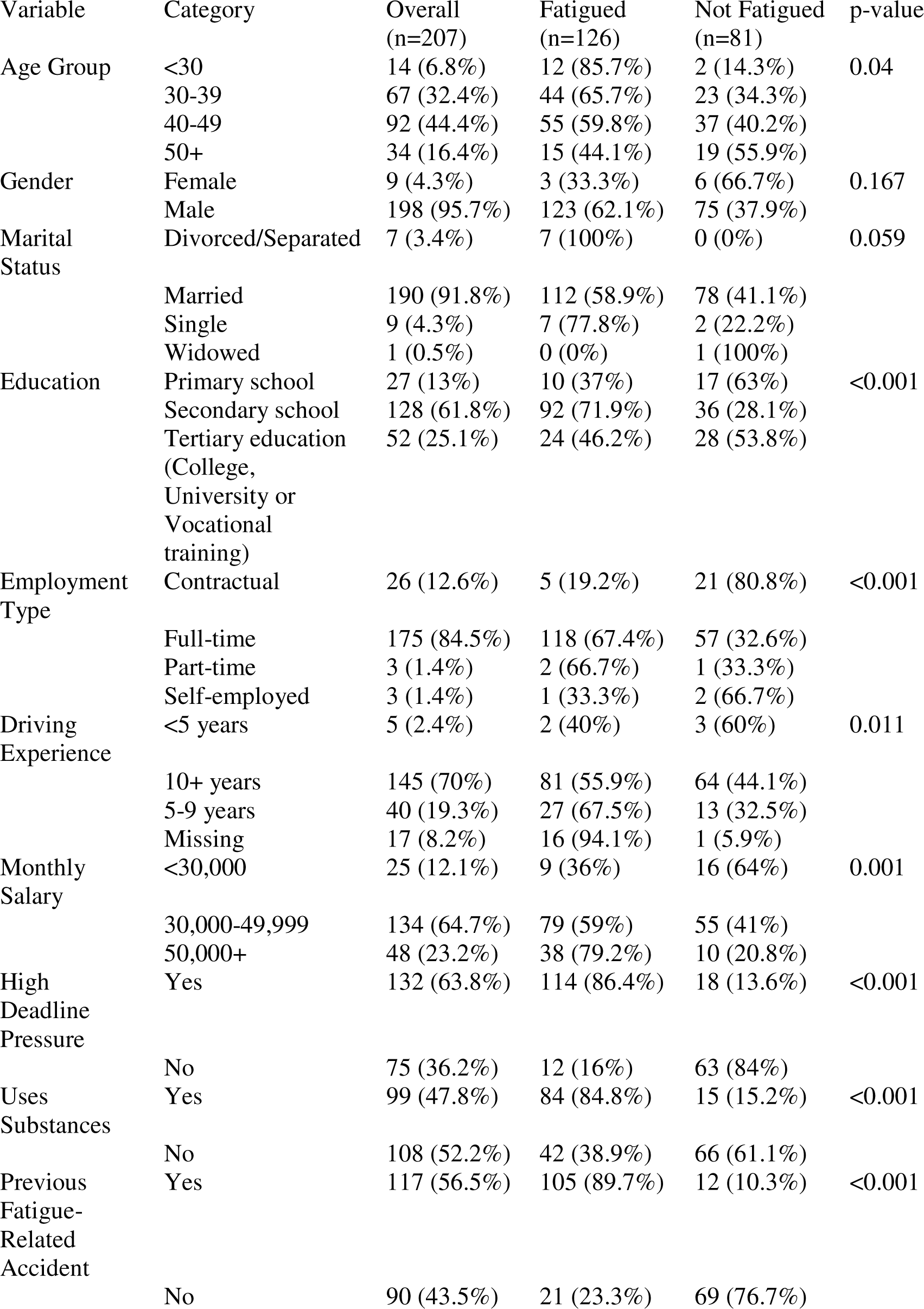
Socio-demographic and Occupational Characteristics of Long-Distance Truck Drivers, Overall and by Fatigue Status.

**Table 2:** Logistic Regression Analysis of Factors Associated with Fatigue in Long-Distance Truck Drivers.

| Variable | Adjusted OR | 95% CI | p-value |
| --- | --- | --- | --- |
| High Deadline Pressure | 20.50 | 1.57 – 31.00 | 0.000*** |
| Substance Use | 14.37 | 1.62 – 26.00 | 0.017* |

The LASSO procedure selected variables for further analysis, as shown in figure 2 and figure 3. When these variables were entered into a multivariable logistic regression model to estimate adjusted Odds Ratios (ORs) and 95% confidence intervals (CIs), only two of them demonstrated statistically significant independent associations with fatigue. As shown in Table 2, high deadline pressure was the strongest identified risk factor. Drivers experiencing high deadline pressure had over twenty times the odds of fatigue compared to those with low pressure (OR = 20.50, 95% CI: 1.57–31.00, p < 0.001). Substance use was also a major and significant predictor; drivers who used substances such as alcohol, khat, or stimulants to manage driving demands had over fourteen times the odds of being classified as fatigued (OR = 14.37, 95% CI: 1.62–26.00, p = 0.017).

**Fig. 1.**
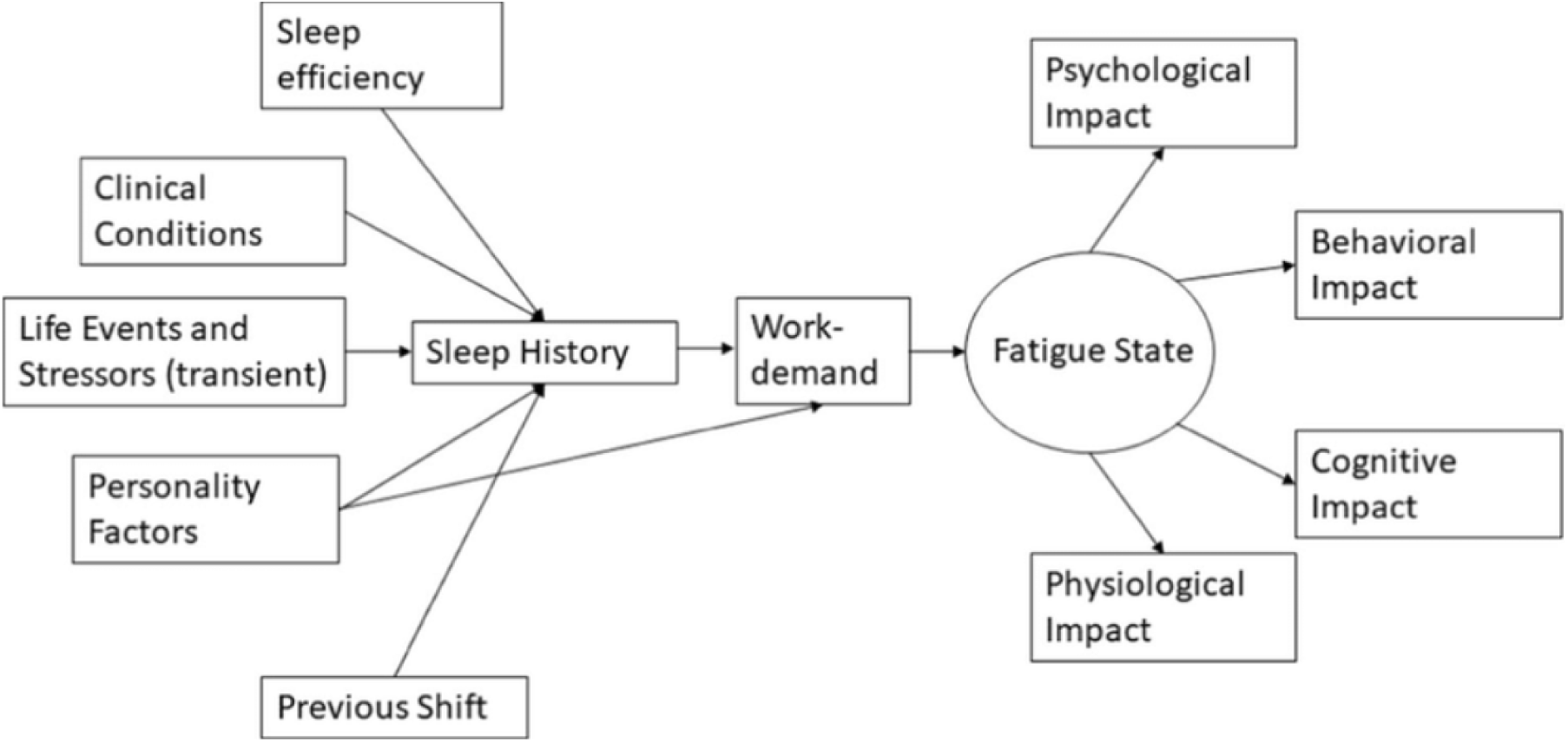
Conceptual framework adopted to understand fatigue in long-distance truck drivers. Source (Drews et al., 2020)

**Figure 2.**
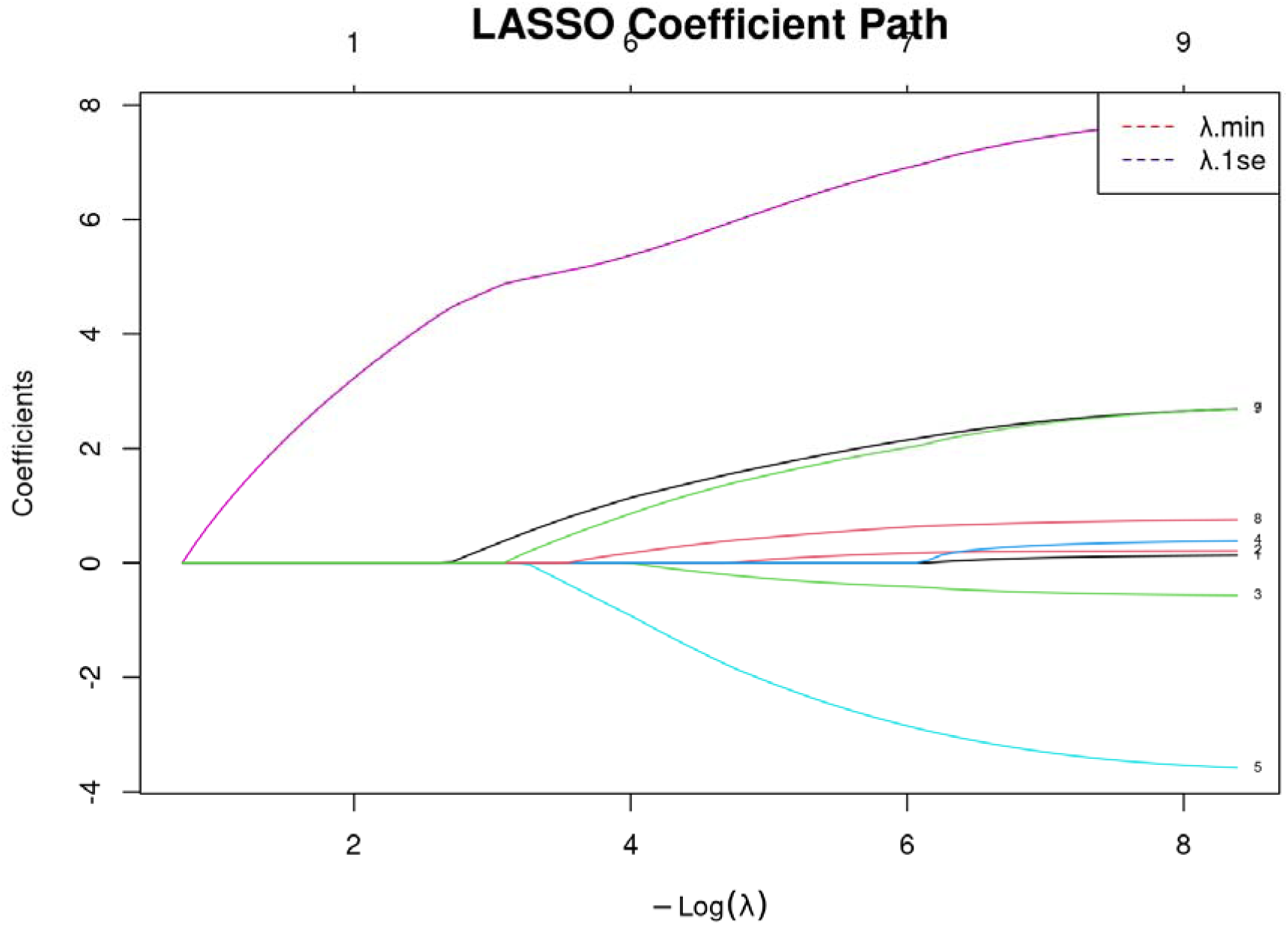
Regularization Paths of Lasso Coefficients on High-Dimensional Data.

**Figure 3.**
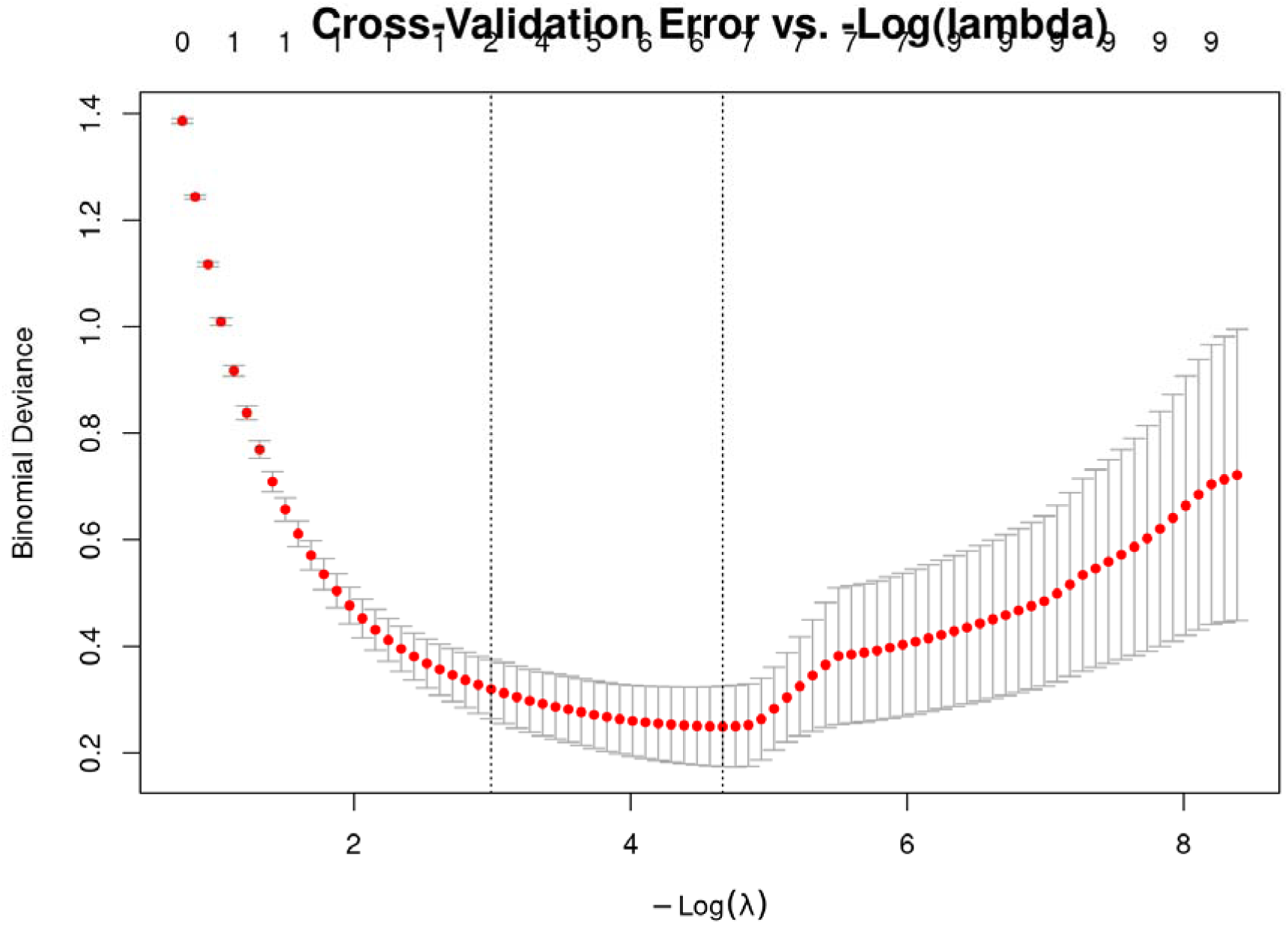
Cross-Validation Mean Squared Error for Lasso Regularization Parameter (λ) Selection.

The final multivariable model, incorporating these two key predictors, exhibited excellent performance. The model’s discriminatory power, measured by the area under the Receiver Operating Characteristic Curve (AUC), was 0.98, indicating excellent ability to distinguish between fatigued and non-fatigued drivers.

To assess the robustness and stability of the model, internal validation was performed using bootstrap resampling with 1,000 iterations. This method involves repeatedly drawing random samples with replacement from the original dataset to create multiple bootstrap samples, each used to refit the model. The performance metrics (AUC, sensitivity, specificity) are then calculated across all iterations to provide a robust estimate of how the model would be expected to perform on new data from the same population.

The bootstrap validation confirmed the model’s excellent and stable performance. The mean Area Under the ROC Curve (AUC) across the 1,000 bootstrap samples was 0.987 (95% CI: 0.983 - 0.991), presented in figure 4 and figure 5, demonstrating consistently high discriminatory power. Similarly, the model maintained high classification accuracy, with a mean sensitivity of 85% and a mean specificity of 86%. This process confirms that the model is not overfitted to the specific sample and is likely to perform well on similar populations of long-distance truck drivers.

**Figure 4.**
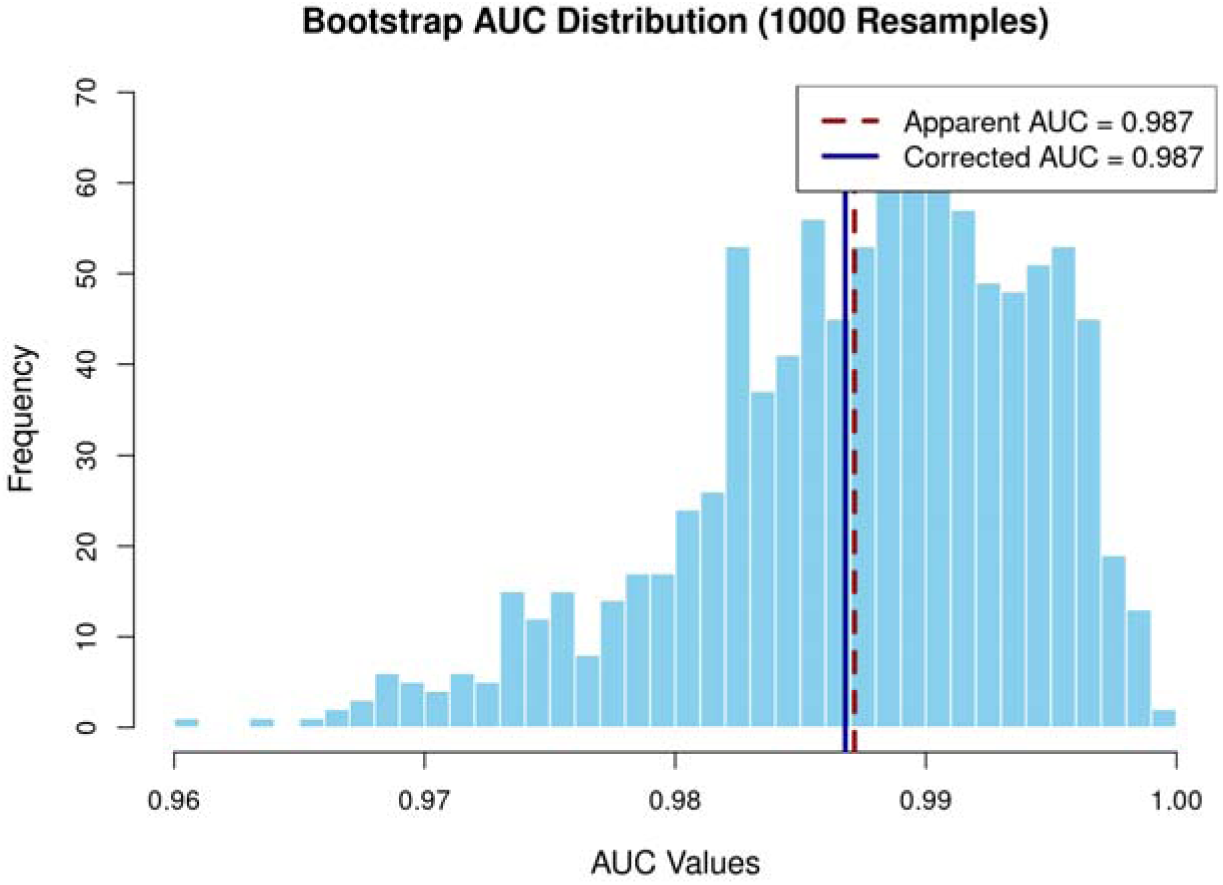
Bootstrap Distribution of the Area Under the ROC Curve (AUC) for the Logistic Regression Model.

This study was conducted in strict adherence to international ethical standards, with informed consent obtained from all participants. Limitations include the potential for recall and social desirability bias in self-reported data, and the cross-sectional design precludes causal inference. However, the use of a validated instrument and statistical safeguards like LASSO regularization and internal validation strengthen the internal validity of the reported associations. The analysis provides confidence that high deadline pressure and substance use are central, significant factors associated with fatigue in this occupational group.

## 4. Discussion

### 4.1. Overview of Principal Findings

This study sought to identify the key determinants of fatigue among long-distance truck drivers in East Africa, a critical public health and road safety issue. Through a rigorous analytical process employing LASSO regression for variable selection and multivariable logistic modeling, we identified two robust, independent predictors of significant fatigue: High Deadline Pressure and Substance Use. The final model exhibited near-perfect discriminatory power (AUC = 0.987) and maintained its robustness upon internal validation. Crucially, the model’s parsimony was achieved by isolating these two potent factors from other potential demographic and occupational confounders, such as a lack of employer-provided medical support or insufficient knowledge of ILO guidelines, which were found to have negligible independent effects. The findings collectively paint a picture of fatigue that is less about individual driver characteristics or even some institutional supports, and more profoundly shaped by intense, immediate work pressures and the maladaptive behavioural coping mechanisms they engender.

### 4.2. Substance Use: A Maladaptive Coping Mechanism

The use of substances such as alcohol, khat, or stimulants was a significant predictor of fatigue, associated with over a fourteen-fold increase in the odds of fatigue (aOR: 14.37). This finding is consistent with a substantial body of literature which identifies stimulant use as a common, albeit dangerous, strategy to extend waking hours and meet demanding delivery schedules (Bakowski et al., 2020; Girotto et al., 2015). In our model, substance use appears not as an isolated behaviour but as a critical mediator in the stress-fatigue pathway. The strong independent effect of substance use, even when considered alongside extreme deadline pressure, suggests it is a primary driver of fatigue in its own right, likely by disrupting natural sleep architecture, leading to poorer quality rest, increased sleep debt, and greater subsequent fatigue, thereby perpetuating the need for further stimulant use (Montoro et al., 2022). The fact that other variables like employer support were non-significant underscores that this coping mechanism persists regardless of the broader occupational safety climate, highlighting its entrenched and potentially individualistic nature.

### 4.3. The Foundational Role of Employer Support Systems

Contrary to expectations, the lack of employer-provided medical support was not retained as a significant independent predictor in the final model. This does not negate the importance of organizational support but suggests that in the high-pressure environment captured by our data, its effect is eclipsed by more immediate, visceral pressures. It is plausible that the pervasive presence of high deadline pressure creates a work culture where the absence of such support is the norm, thereby neutralizing its potential as a differentiating protective factor. This finding aligns with occupational health models that posit organizational support as a buffer against burnout (Arnold et al., 1997; Chen & Zhang, 2016); its non-significance here points to a sector where this buffer has been completely overwhelmed by operational demands.

### 4.4. Knowledge as Power: The Protective Effect of ILO Awareness

Our final model indicated that insufficient knowledge of ILO regulations was not a significant independent predictor of fatigue. This is a critical finding, as it suggests that in an environment defined by intense commercial pressure, knowledge alone may not be power. Drivers who are aware of their rights to adequate rest may lack the agency to refuse unsafe schedules due to economic precarity or fear of reprisal. The dominance of high deadline pressure as a predictor implies that even knowledgeable drivers are often compelled to violate guidelines to meet their contractual obligations. This result resonates with health behaviour models, where knowledge is a prerequisite for action (Dorrian et al., 2006; Garz et al., 2021); however, it suggests that without the agency to act, knowledge’s protective effect is nullified by overwhelming pressure.

### 4.5. Deconstructing the Paradox: Work-Rest Records as a Marker of Safety Climate

The variable for poor work-rest record-keeping was not selected in the final model. Its absence is informative. While formal record-keeping can be an indicator of a safety-compliant company (Casey et al., 2024; Cori et al., 2021), its non-significance in the presence of high deadline pressure suggests that in this context, such records may be symbolic rather than substantive. A company may maintain records as a formality while simultaneously enforcing schedules that make adherence to work-rest rules impossible. Therefore, the predictive power lies not in the paperwork, but in the tangible, enforced operational pressure. The model reveals that it is the direct experience of high deadline pressure, rather than the proxy of record-keeping, that is the true driver of fatigue outcomes.

### 4.6. Synthesis and Contribution to the Literature

When synthesized, our findings present a compelling and stark narrative. The typical long-distance truck driver in our East African cohort operates within a system where intense, unrelenting deadline pressure is the primary determinant of their fatigue state. This pressure directly increases fatigue and also manifests indirectly by fostering the use of stimulants as a coping mechanism, which in turn exacerbates the problem. The strong performance of our model (AUC = 0.987), which excluded variables like employer support and ILO knowledge, suggests that in this specific high-stakes environment, these systemic factors are secondary to the immediate economic and operational drivers. This study makes a significant contribution by highlighting that while supportive structures and knowledge are important, interventions that do not first address the root cause of excessive deadline pressure and its consequent substance use are unlikely to succeed.

### 4.7. Strengths and Limitations

The major strength of this study lies in its rigorous methodology. The application of LASSO regression effectively mitigated the risks of overfitting and multicollinearity, leading to a parsimonious and stable model containing only the two most potent predictors. The model’s exceptional discriminatory power (AUC = 0.987) and successful internal validation confirm that its performance is not an artefact of the specific sample but is generalizable to similar populations. However, several limitations must be acknowledged. The cross-sectional nature precludes definitive causal inference; while it is logical to assume that pressure and substance use cause fatigue, reverse causality where fatigued drivers turn to substances to cope is also plausible. The reliance on self-reported data introduces potential for social desirability and recall bias. Despite these limitations, the statistical safeguards employed and the remarkable strength of the associations provide considerable confidence in the internal validity of the results.

### 4.8. Implications for Policy and Practice

The findings from this study yield clear, actionable recommendations:

i. **For Policymakers and Regulators:** There is an urgent need to bridge the gap between formal ILO standards and on-the-ground practice. This can be achieved by mandating and enforcing driver and employer education on hours-of-service regulations and integrating fatigue management principles into national road safety strategies.
ii. **For Trucking Companies and Employers:** A paradigm shift from viewing drivers as expendable assets to valuing them as core human capital is required. Employers should institutionalize comprehensive driver wellness programs that include: (1) mandatory, confidential medical support and health screenings; (2) strict enforcement of work-rest schedules within a transparent record-keeping system; and (3) a strict, supportive policy on substance use, focusing on rehabilitation over punishment.
iii. **For Future Research:** Longitudinal studies are needed to establish causal pathways and to evaluate the real-world impact of interventions based on these findings. Qualitative research would be invaluable for deepening our understanding of the drivers’ lived experiences, the barriers to adopting safe practices, and the dynamics of substance use within this community.

### 4.9. Conclusion

In conclusion, this study demonstrates that fatigue among long-distance truck drivers in East Africa is a multifactorial condition rooted in a complex interplay of behavioural risk and systemic failure. The potent roles of substance use, lack of medical support, insufficient regulatory knowledge, and poor work-rest governance highlight that solutions must extend beyond the individual driver to encompass the organizational and regulatory structures that shape their working environment. Tackling this pervasive issue requires a concerted, multi-stakeholder approach focused on building a robust safety culture, enhancing driver welfare, and rigorously applying international labour standards to protect the health of drivers and the safety of all road users.

## Supporting information

Ethical approval

STROBE checklist

## Data Availability

The datasets generated and analyzed during the current study are not publicly available due to ethical and confidentiality considerations but are available from the corresponding author upon reasonable request.

Supplementary materials, including the anonymized dataset, R analysis scripts (including LASSO and bootstrap procedures), model output files, additional diagnostic figures, and the structured study questionnaire, are available upon request for purposes of academic verification and replication, subject to institutional ethical approval requirements.

## Acknowledgements

We acknowledge the valuable contributions of Dr. Nelson Kilimo, Dr. Kellen Karimi, and Dr. Verena Struckmann to the study’s design, methodological approach, and data analysis. We are also deeply thankful to Dr. Olipher Makwaga at the Kenya Medical Research Institute (KEMRI) in Busia for her invaluable support in guiding with manuscript development. We extend our appreciation to the dedicated team of research assistants who conducted the surveys and interviews at various truck stops and border crossings across East Africa. This work would not have been possible without the participation of the long-distance truck drivers who generously shared their time and experiences.

## References

1. Abd-elnaby, M., Alfonse, M., & Roushdy, M. (2022). A Hybrid Mutual Information-LASSO-Genetic Algorithm Selection Approach for Classifying Breast Cancer. In D. A. Magdi, Y. K. Helmy, M. Mamdouh, & A. Joshi (Eds.), Digital Transformation Technology (Vol. 224, pp. 547–560). Springer Singapore. 10.1007/978-981-16-2275-5_36

2. Ahlström, C., & Anund, A. (2025). Development of sleepiness in professional truck drivers: Real road testing for driver drowsiness and attention warning ( DDAW ) system evaluation. Journal of Sleep Research, 34(2), e14259. 10.1111/jsr.14259

3. Akyurek, G., Kaya Ozturk, L., & Gurlek, S. (2025). Investigation of factors predicting quality of life of drivers working on long-haul transport: Pain, fatigue, stress and work role functions. International Journal of Occupational Safety and Ergonomics, 1–7. 10.1080/10803548.2025.2474835

4. Al-Mekhlafi, A.-B. A., Isha, A. S. N., Chileshe, N., Abdulrab, M., Kineber, A. F., & Ajmal, M. (2021). Impact of Safety Culture Implementation on Driving Performance among Oil and Gas Tanker Drivers: A Partial Least Squares Structural Equation Modelling (PLS-SEM) Approach. Sustainability, 13(16), 8886. 10.3390/su13168886

5. Al-Mekhlafi, A.-B. A., Isha, A. S. N., Chileshe, N., Abdulrab, M., Saeed, A. A. H., & Kineber, A. F. (2021). Modelling the Relationship between the Nature of Work Factors and Driving Performance Mediating by Role of Fatigue. International Journal of Environmental Research and Public Health, 18(13), 6752. 10.3390/ijerph18136752

6. Amoadu, M., Ansah, E. W., & Sarfo, J. O. (2023). Psychosocial work factors, road traffic accidents and risky driving behaviours in low-and middle-income countries: A scoping review. IATSS Research, 47(2), 240–250. 10.1016/j.iatssr.2023.03.005

7. Arnold, P. K., Hartley, L. R., Corry, A., Hochstadt, D., Penna, F., & Feyer, A. M. (1997). Hours of work, and perceptions of fatigue among truck drivers. Accident Analysis & Prevention, 29(4), 471–477. 10.1016/S0001-4575(97)00026-2

8. Bakowski, A., Jurecki, R., Radziszewski, L., & Swietlik, P. (2020). The analysis of the relations between road vehicle traffic parameters and the number of road accidents in subsequent hours of the day—A case study. 2020 XII International Science-Technical Conference AUTOMOTIVE SAFETY, 1–7. 10.1109/AUTOMOTIVESAFETY47494.2020.9293511

9. Barck-Holst, P., Nilsonne, Å., Åkerstedt, T., & Hellgren, C. (2021). Coping with stressful situations in social work before and after reduced working hours, a mixed-methods study. European Journal of Social Work, 24(1), 94–108. 10.1080/13691457.2019.1656171

10. Baulk, S. D., & Fletcher, A. (2012). At home and away: Measuring the sleep of Australian truck drivers. Accident Analysis & Prevention, 45, 36–40. 10.1016/j.aap.2011.09.023

11. Belzer, M. H., & Sedo, S. A. (2018). Why do long distance truck drivers work extremely long hours? The Economic and Labour Relations Review, 29(1), 59–79. 10.1177/1035304617728440

12. Bener, A., Yildirim, E., Özkan, T., & Lajunen, T. (2017). Driver sleepiness, fatigue, careless behavior and risk of motor vehicle crash and injury: Population based case and control study. Journal of Traffic and Transportation Engineering (English Edition), 4(5), 496–502. 10.1016/j.jtte.2017.07.005

13. Casey, G. J., Miles-Johnson, T., & Stevens, G. J. (2024). Heavy vehicle driver fatigue: Observing work and rest behaviours of truck drivers in Australia. Transportation Research Part F: Traffic Psychology and Behaviour, 104, 136–153. 10.1016/j.trf.2024.05.016

14. Chalder, T., Berelowitz, G., Pawlikowska, T., Watts, L., Wessely, S., Wright, D., & Wallace, E. P. (1993). Development of a fatigue scale. Journal of Psychosomatic Research, 37(2), 147–153. 10.1016/0022-3999(93)90081-P

15. Chan, M. (2011). Fatigue: The most critical accident risk in oil and gas construction. Construction Management and Economics, 29(4), 341–353. 10.1080/01446193.2010.545993

16. Charan, J., & Biswas, T. (2013). How to Calculate Sample Size for Different Study Designs in Medical Research? Indian Journal of Psychological Medicine, 35(2), 121–126. 10.4103/0253-7176.116232

17. Chen, C., & Zhang, J. (2016). Exploring background risk factors for fatigue crashes involving truck drivers on regional roadway networks: A case control study in Jiangxi and Shaanxi, China. SpringerPlus, 5(1), 582. 10.1186/s40064-016-2261-y

18. Cori, J. M., Downey, L. A., Sletten, T. L., Beatty, C. J., Shiferaw, B. A., Soleimanloo, S. S., Turner, S., Naqvi, A., Barnes, M., Kuo, J., Lenné, M. G., Anderson, C., Tucker, A. J., Wolkow, A. P., Clark, A., Rajaratnam, S. M. W., & Howard, M. E. (2021). The impact of 7-hour and 11-hour rest breaks between shifts on heavy vehicle truck drivers’ sleep, alertness and naturalistic driving performance. Accident Analysis & Prevention, 159, 106224. 10.1016/j.aap.2021.106224

19. Coutinho, K. B. (n.d.). Monitoring real time data of the seafarer work and rest hours onboard merchant ships &stakeholder utilization of the data to manage operations risk in merchant shipping.

20. Dorrian, J., Lamond, N., Van Den Heuvel, C., Pincombe, J., Rogers, A. E., & Dawson, D. (2006). A Pilot Study of the Safety Implications of Australian Nurses’ Sleep and Work Hours. Chronobiology International, 23(6), 1149–1163. 10.1080/07420520601059615

21. Drews, F. A., Rogers, W. P., Talebi, E., & Lee, S. (2020). The Experience and Management of Fatigue: A Study of Mine Haulage Operators. Mining, Metallurgy & Exploration, 37(6), 1837–1846. 10.1007/s42461-020-00259-w

22. El_Rahman, S. A. (2021). Predicting breast cancer survivability based on machine learning and features selection algorithms: A comparative study. Journal of Ambient Intelligence and Humanized Computing, 12(8), 8585–8623. 10.1007/s12652-020-02590-y

23. Faul, F., Erdfelder, E., Buchner, A., & Lang, A.-G. (2009). Statistical power analyses using G*Power 3.1: Tests for correlation and regression analyses. Behavior Research Methods, 41(4), 1149–1160. 10.3758/BRM.41.4.1149

24. Friswell, R., & Williamson, A. (2013). Comparison of the fatigue experiences of short haul light and long distance heavy vehicle drivers. Safety Science, 57, 203–213. 10.1016/j.ssci.2013.02.014

25. Garz, S., Giné, X., Karlan, D., Mazer, R., Sanford, C., & Zinman, J. (2021). Consumer Protection for Financial Inclusion in Low and Middle Income Countries: Bridging Regulator and Academic Perspectives (SSRN Scholarly Paper No. 3750236). 10.2139/ssrn.3750236

26. Girotto, E., de Andrade, S. M., Mesas, A. E., González, A. D., & Guidoni, C. M. (2015). Working conditions and illicit psychoactive substance use among truck drivers in Brazil. Occupational and Environmental Medicine, 72(11), 764–769.

27. Hamid, M., Samuel, S., Borowsky, A., Horrey, W. J., & Fisher, D. L. (2016). Evaluation of Training Interventions to Mitigate Effects of Fatigue and Sleepiness on Driving Performance. Transportation Research Record: Journal of the Transportation Research Board, 2584(1), 30–38. 10.3141/2584-05

28. Hanowski, R. J., Hickman, J., Fumero, M. C., Olson, R. L., & Dingus, T. A. (2007). The sleep of commercial vehicle drivers under the 2003 hours-of-service regulations. Accident Analysis & Prevention, 39(6), 1140–1145. 10.1016/j.aap.2007.02.011

29. Hasan, N. A. C., Karuppiah, K., Hamzah, N. A., Juzad, K. M., & Tamrin, S. B. M. (2022). PREVALENCE OF DRIVING FATIGUE AND ITS ASSOCIATED FACTORS AMONG LOGISTIC TRUCK DRIVERS IN MALAYSIA. Malaysian Journal of Public Health Medicine, 22(3), Article 3. 10.37268/mjphm/vol.22/no.3/art.1745

30. Jackson, C. (2015). The Chalder Fatigue Scale (CFQ 11). Occupational Medicine, 65(1), 86–86. 10.1093/occmed/kqu168

31. Li, Y., Lu, F., & Yin, Y. (2022). Applying logistic LASSO regression for the diagnosis of atypical Crohn’s disease. Scientific Reports, 12(1), 11340. 10.1038/s41598-022-15609-5

32. Maldonado, C., Mitchell, D., SR, T., & Driver, H. (2002). Sleep, work schedules and accident risk in South African long-haul truck drivers. South African Journal of Science, 98, 319–324.

33. Montoro, L., Cendales, B., Alonso, F., Gonzalez-Marin, A., Lijarcio, I., Llamazares, J., & Useche, S. A. (2022). Essential…but also vulnerable? Work intensification, effort/reward imbalance, fatigue and psychological health of Spanish cargo drivers during the COVID-19 pandemic. PeerJ, 10, e13050. 10.7717/peerj.13050

34. Omondiagbe, D. A., Veeramani, S., & Sidhu, A. S. (2019). Machine Learning Classification Techniques for Breast Cancer Diagnosis. IOP Conference Series: Materials Science and Engineering, 495, 012033. 10.1088/1757-899X/495/1/012033

35. Philip, P., Taillard, J., & Micoulaud-Franchi, J.-A. (2019). Sleep Restriction, Sleep Hygiene, and Driving Safety. Sleep Medicine Clinics, 14(4), 407–412. 10.1016/j.jsmc.2019.07.002

36. Robin, X., Turck, N., Hainard, A., Tiberti, N., Lisacek, F., Sanchez, J.-C., & Müller, M. (2011). pROC: An open-source package for R and S+ to analyze and compare ROC curves. BMC Bioinformatics, 12(1), 77. 10.1186/1471-2105-12-77

37. Rodríguez Del Águila, M., & González-Ramírez, A. (2014). Sample size calculation. Allergologia et Immunopathologia, 42(5), 485–492. 10.1016/j.aller.2013.03.008

38. Soliani, R. D., Silva, L. B. D., & Barbosa, A. D. S. (2023). THE EFFECTS OF FATIGUE ON TRUCK DRIVERS IN CARGO TRANSPORTATION: A LITERATURE REVIEW. 48.

