## Supplementary material for "Identifying the Key Predictors of Occupational Fatigue among Long-Distance Truck Drivers in East Africa: A LASSO-Regularized Regression Approach": Ethical approval

UNIVERSITY OF NAIROBI

**UNIVERSITY OF NAIROBI**
**FACULTY OF HEALTH SCIENCES**
**P O BOX 19676 Code 00202**
**Telegrams: varsity**
**(254-020) 2726300**

**KNH-UoN ERC**
****
**Website: http://www.erc.uonbi.ac.ke**
**Facebook: https://www.facebook.com/uonknh.erc**
**Twitter: @UONKNH_ERC https://twitter.com/UONKNH_ERCs**

**KENYATTA NATIONAL HOSPITAL**
**P O BOX 20723 Code 00202**
**Fax: 725272**
**Telegrams: MEDSUP, Nairobi**

Ref: KNH-ERC/RR/804

26th September, 2024

Nelson Kiptoo Kilimo
Reg. No. H57/42656/2022
Dept. of Public and Global Health
Faculty of Health Sciences
University of Nairobi

Dear Nelson,

**Research Proposal: Prevalence and risk factors for occupational fatigue among long-distance truck drivers at Kenyan-Ugandan border (P555/07/2024)**

---

This is to acknowledge receipt of your research proposal and to inform you that upon review, the KNH- UoN Ethics and Research Committee made the following observations and suggestions:

*Preliminaries*

1. Study Title: Replace the phrase "*…at Kenya-Ugandan border*", with "*…**at the Kenya-Uganda border***".
2. Abstract:
    i. Provide a brief explanation of how data will be analyzed.
    ii. Summarize and combine the subsections 'Results, Discussion and Conclusion' under the subtitle "***Expected Outcomes***".

*Chapter Three- Research Methodology*

3. Section 3.5.1, Inclusion Criteria: State the rationale for using 28 years as the cut-off age for study participants.
4. Include a section on Study Limitations and Mitigation Measures. Ensure to state the study limitations and describe the measures that will be undertaken to minimize their effects.
5. Data Analysis: Attach dummy illustrations/ tables for data presentation. These can be presented as appendices.

*Appendices*

6. Informed Consent Form:
    i. Study Procedure- State the estimated time the participants will take to complete the questionnaire.
    ii. Provide contact details of your supervisors and those of the KNH- UoN ERC.
    iii. Include the 'Participant's Statement' and 'Researcher's Statement'.
    iv. Two informed consent forms are attached. Which one will you use?
7. Study Questionnaire: Collect data on age and salary as continuous variables. Categorization should be done during data analysis.

**Recommendation**

Revise and resubmit **three (3)** copies of the full proposal inclusive of the Application Form within a period of **four (4)** weeks with effect from the date of this letter. Include a cover letter that summarizes how you have addressed the comments and note the page number(s) where the changes have been made.

This can be presented in form of a table that summarizes how all the issues raised have been addressed and further highlight/ bold the corrections in body of the proposal for ease of reference.

Yours sincerely,

**PROF. BEATRICE K.M. AMUGUNE**
**SECRETARY, KNH- UoN ERC**

c.c. The Dean, Faculty of Health Sciences, UoN
The Senior Director, CS, KNH
The Chairperson, KNH- UoN ERC
The Chair, Dept. of Public and Global Health, UoN
Supervisors: Dr. Kellen Karimi, Dept. of Public and Global Health, UoN
Dr. Verena Struckmann, Dept. of Healthcare Management, Technical University of Berlin, Germany
