## Supplementary material for "Identifying the Key Predictors of Occupational Fatigue among Long-Distance Truck Drivers in East Africa: A LASSO-Regularized Regression Approach": STROBE checklist

STROBE Statement—checklist of items that should be included in reports of observational studies

|  | Item No | Recommendation |
| --- | --- | --- |
| **Title and abstract** | 1 | (*a*) This was an *analytical cross-sectional study*. |
|  |  | (*b*) Abstract clearly states objectives, design, sample, measures, analysis (LASSO + logistic regression), results (prevalence and predictors), and conclusions (page 1). |
| Introduction | | |
| Background/rationale | 2 | The Introduction explains the growing burden of truck-driver fatigue globally and the scarcity of evidence for East African transport corridors, establishing the need for the investigation (page 3). |
| Objectives | 3 | The study aimed to determine the **prevalence** of occupational fatigue and identify **key predictors** among long-distance truck drivers; hypotheses implied that organizational and behavioural factors increase fatigue risk (page 4). |
| Methods | | |
| Study design | 4 | An analytical cross-sectional design is stated clearly at the beginning of the Methods section (page 5). |
| Setting | 5 | The setting is Busia and Malaba border points on the Kenya-Uganda corridor. Relevant dates: data collection occurred during 3 week survey period from January 6^th^ to January 24^th^ 2025 (page 5). |
| Participants | 6 | (*a*) *Cross-sectional study*— Eligibility: long-distance truck drivers aged ≥21years, actively engaged in cross-border freight operations, present at Busia/Malaba during data collection; Departed from Mombasa sea port. Sampling: Systematic random sampling of every 2^nd^ driver at truck stops. Exclusively male participants (page 5). |
|  |  | (*b*) *N/A* |
| Variables | 7 | Fatigue outcome: measured using the validated **Chalder Fatigue Scale.** Predictors: demographics, work patterns, shift length, stimulant use, sleep habits, pressure to meet deadlines, company shift policies. Confounders considered: age, experience, rest quality (page 5). |
| Data sources/ measurement | 8* | All variables were collected using **structured interviewer-administered questionnaires**. Fatigue assessed per Chalder Fatigue Scale scoring guidelines. Exposure and confounder variables self-reported. Single group only - no differential measurement (page 5). |
| Bias | 9 | Efforts included standardized data collection procedures, training of research assistants, validated fatigue scale, and use of LASSO to reduce multicollinearity and overfitting. Potential recall and self-report bias acknowledged (page 6). |
| Study size | 10 | The Kelsey et al. formula was adopted to determine the a priori sample size. Assuming a 95% confidence level, 80% power, and an expected OR of 2.0, with exposure proportions of 23% (exposed) and 12.9% (unexposed), a minimum total sample size of 207 participants was derived (page 5). |
| Quantitative variables | 11 | Continuous variables e.g., shift length, retained or categorized based on theoretical relevance; LASSO performed automatic shrinkage/selection to correct for sparse data (page 5). |
| Statistical methods | 12 | (*a*) Descriptive statistics summarized characteristics. LASSO with 10-fold CV selected predictors. Selected variables entered into multivariable logistic regression to estimate **adjusted odds ratios** (page 6). |
|  |  | (*b*) No subgroup or interaction analyses planned due to analytical focus on predictor selection (page 6). |
|  |  | (*c*) Missing data were minimal. The missing observations were managed using complete-case analysis, which was deemed appropriate given the minimal missingness. Furthermore, any remaining missing or erroneously wild continuous values were addressed by imputing them with the median of their respective distributions (page 6). |
|  |  | (*d*) *N/A* |
|  |  | (*e*) Bootstrap validation (1,000 samples) assessed stability of the model and internal validity (page 6). |

Continued on next page

| Results | | |
| --- | --- | --- |
| Participants | 13* | (a) Flow: Drivers approached (221) → Screened (221) → Consented (211) → Filled questionnaire (211) → Included in final analysis (207). Total analysed: 207 (page 6). |
|  |  | (b) A total of 221 drivers were approached and screened for the study. Of these, 10 drivers did not consent to participate, leaving 211 consenting participants. However, during the data collection process, four participants stopped midway and were subsequently excluded from the final analysis due to the urgent need to advance in the truck queue and clear with the customs office. Consequently, the final analysis included 207 drivers (page 6). |
|  |  | (c) N/A |
| Descriptive data | 14* | (a) A total of 207 mostly male (96%) long-distance drivers participated, with a median age of 42. Most were married, educated to secondary level, and full-time employed. They earned a median KSh 35,000, supported five dependents, had 14 years’ driving experience, and typically drove long daily hours (page 6). |
|  |  | (b) No variable had missing data; however, several extreme outliers (e.g., salary 4,000,000 KSh, age 5600, household dependents 900) were identified and imputed using median values to maintain data integrity (page 6). |
|  |  | (c) *Not applicable* |
| Outcome data | 15* | *Cross-sectional study -* Reported numbers of outcome events or summary measures (page 6). |
| Main results | 16 | (*a*) Univariable models showed strong associations between fatigue and deadline pressure, driving hours, overnight stays, and substance use, with precise 95% CIs. The multivariable model adjusted for all significant univariable predictors to address confounding. Adjusted odds remained high for these occupational and behavioural factors (page 6). |
|  |  | (*b*) Continuous variables were categorised using predefined boundaries: household dependents (0–3 small, 4–7 medium, >7 large), and fatigue status based on the Chalder Fatigue Scale cut-off. All other continuous predictors remained in their original scale to preserve information (page 6). |
|  |  | (*c*) Not applicable |
| Other analyses | 17 | Stability assessed using **bootstrap validation** (AUC=0.987). No subgroup or interaction analysis performed (page 7, page 8, and page 9). |
| Discussion | | |
| Key results | 18 | Summarized relative to objectives: high prevalence of fatigue and key predictors identified (page 10). |
| Limitations | 19 | Self-report bias, cross-sectional design (no causal inference), potential unmeasured confounding, and corridor-based recruitment limiting generalizability to all East African drivers (page 4). |
| Interpretation | 20 | Cautious interpretation given design limitations; results align with similar research; LASSO improves robustness of predictor selection (page 11). |
| Generalisability | 21 | Findings generalizable to similar East African long-distance truck corridors with comparable work conditions; context-specific limitations acknowledged (page 12). |
| Other information | | |
| Funding | 22 | No external funding received. |

*Give information separately for cases and controls in case-control studies and, if applicable, for exposed and unexposed groups in cohort and cross-sectional studies.

**Note:** An Explanation and Elaboration article discusses each checklist item and gives methodological background and published examples of transparent reporting. The STROBE checklist is best used in conjunction with this article (freely available on the Web sites of PLoS Medicine at http://www.plosmedicine.org/, Annals of Internal Medicine at http://www.annals.org/, and Epidemiology at http://www.epidem.com/). Information on the STROBE Initiative is available at www.strobe-statement.org.
